# Waist Circumference as a potential indicator of cardiac autonomic profile in Metabolic Syndrome - A cross-sectional study

**DOI:** 10.1101/2023.11.15.23298569

**Authors:** Ankith Bhasi, Oshin Puri, Monika Pathania, Sarama Saha, Meenakshi Dhar, Poorvi Kulshreshta

**Affiliations:** Department of Medicine, All India Institute of Medical Sciences, Rishikesh, Uttarakhand-249203, India; MBBS Intern/MS5, All India Institute of Medical Sciences, Rishikesh, Uttarakhand-249203, India; Additional Professor, Department of Geriatric Medicine, All India Institute of Medical Sciences, Rishikesh, Uttarakhand-249203, India; Associate Professor, Department of Biochemistry, All India Institute of Medical Sciences, Rishikesh, Uttarakhand-249203, India; Additional Professor and Head, Department of Geriatric Medicine, All India Institute of Medical Sciences, Rishikesh, Uttarakhand-249203, India; Additional Professor, Department of Physiology, All India Institute of Medical Sciences, Rishikesh, Uttarakhand-249203, India

**Keywords:** Metabolic Syndrome. Waist circumference, Heart Rate Variability, Obesity

## Abstract

**Introduction:** - Metabolic Syndrome (MetS), a global epidemic on the rise, affects about one-third of the world’s population. MetS poses significant cardiovascular risk often assessed through the cardiac autonomic function test Heart Rate Variability (HRV). This study attempts to identify a suitable anthropometric or biochemical proxy for resource-intensive HRV to assess cardiac autonomic profile in primary and community healthcare setups.

**Methodology:** - This cross-section study assessed relevant anthropometric, biochemical, and HRV parameters of 174 patients recently diagnosed with MetS.

**Results:** - Waist circumference (WC) had a significant (p<0.05) moderate to strong (r=0.49) positive correlation with sympathetic predominant HRV parameters - LFnu and LF/HF ratio. Parasympathetically modulated HRV parameters like Average RR (r=-0.17), rMSSD (r=-0.23), and HF (r=-0.49) had a significant (p<0.05) negative correlation with WC.

**Conclusion:** - The study identifies WC as a suitable indicator for cardiac autonomic profile in patients with MetS and by extension of cardiovascular posed by the disease.

## Introduction

**-**The prevalence of Metabolic Syndrome (MetS) (as per the National Cholesterol Education Program (NCEP) Adult Treatment Panel (ATP) III definition) among rural and urban Indian populations is 24.6% and 16.9% respectively. (1–3) Even such high numbers are surpassed by many other countries worldwide such as 49% in urban Pakistan, 37.1% in Malaysia, 35.8% in Australia, and 32.8% in Mongolia. (3–7) The International Diabetes Federation (IDF) and NCEP ATP III define MetS as a cluster of inter-connected metabolic abnormalities involving glucose and lipid metabolism, elevated blood pressure, and central obesity. (8) Insulin resistance, atherosclerotic cardiovascular diseases, a general pro-thrombotic and pro-inflammatory state, non-alcoholic steatohepatitis, and reproductive disorders are some of the many comorbidities associated with MetS. (3, 9-13)

Among many, the most well-established and concerning co-morbidity associated with MetS is its cardiovascular risk. Along with hypertension and dyslipidemia, which are components of the disease itself, MetS also creates a pro-thrombotic, pro-inflammatory internal environment and increases circulating reactive oxygen species, all contributing to increased cardiovascular risk among MetS patients. (11, 12, 14) These inter-dependent and co-existing metabolic derangements and the uncertainty of a causal relation between them pose a challenge to understanding cardiovascular risk of MetS. What is the cardiovascular risk size or proportion of elevated glucose, cholesterol, or blood pressure in the background of MetS? Is the cardiovascular risk posed by MetS a summation of the risk posed by its constituent pathological components? If not, is it less or more than the summation? and Why?

Such physiological and pathological systems are a challenge to understand in terms of mathematical summations or other formulations. Moreover, these parameters are evaluated using biochemicals with high time-to-time variability and can be controlled with medication. Once controlled over medication it is tough to establish the cardiovascular damage their derangements might have already done. In an attempt to simplify the complex interplay between MetS components and their resultant cardiovascular risk, Grassi G. et. al. hypothesized that sympathetic overdrive might be a common pathogenic background for all MetS derangements and the cardiovascular risk associated with it. (15)

There is enough evidence suggesting that sympathetic overdrive is associated with MetS as a whole and not just its component metabolic derangements. (16–18) Sympathetic overdrive contributes to the cardiovascular risk associated with MetS and might be a reliable indicator of the cumulative cardiovascular risk of the disease. (15, 19, 20) Cardiac (primarily left ventricular) hypertrophy, and abnormalities in arterial wall thickness, distensibility, and compliance have been demonstrated to stem from this hyperadrenergic pathogenic background of MetS. (15, 21-26) Hence, the autonomic profile especially the cardiac autonomic profile might be a suitable indicator of cardiovascular risk in MetS.

The activity of the autonomic nervous system can be evaluated through Autonomic Function Tests (AFTs) such as Heart Rate Variability (HRV), Blood Pressure Variability, Cold Pressor Test, Valsalva Maneuver, and Galvanic Skin Response to name a few. HRV is often referred to as a cardiac AFT (cAFT) since along with the general autonomic profile, it is a particularly accurate predictor of the autonomic regulation of the heart. (27) AFTs are resource-intensive diagnostics and the equipment as well as technically skilled manpower are seldom available at primary, community, or even district healthcare centers. Hence, even though it is reliable it might have limited practical use at a large scale.

We are curious if a biochemical or anthropometric measure has a significant correlation with the cardiac autonomic profile of MetS to be used as a suitable mass-scale proxy for cAFTs and by extension a determinant of cardiovascular risk. Thus, the current study assesses the correlation between cardiac autonomic function (namely HRV), and biochemical and anthropometric measures of patients with MetS in search of a reliable biochemical or anthropometric proxy for the cardiac autonomic function profile and by extension cardiovascular risk.

## Methodology

### Study Design

This study was conducted as a cross-sectional study.

### Consent and Ethical Considerations

A written well-informed consent was taken from all participants. The study was conducted with due ethical clearance from the Institutional Ethics Committee (IEC). The study complies with the Declaration of Helsinki.

### Study Population

The study was conducted with 174 adults (age > 18 years) who reported to a tertiary healthcare center’s Lifestyle Disease Outpatient Clinic in Uttarakhand, India, and were diagnosed with MetS. Potential participants with liver disease, renal disorder (stage 4 and 5 CKD), hypothyroidism, and suffering from acute illnesses at the time of the study and known cases of MetS were excluded.

### Study Tools and Outcome Measures

#### National Cholesterol Education Program (NCEP) Adult Treatment Panel (ATP) III Definition of Metabolic Syndrome

MetS suspects were diagnosed and hence approached for participation as per the NCEP ATP III definition with abdominal circumference corrected based on IDF criteria for Asians. (28, 29, Table-1)

**Table 1.** NCEP ATP III Diagnostic definition of MetS.

| MetS Component | NCEP-ATP III (2001): 3 of the following 5 parameters. |
| --- | --- |
| Abdominal Circumference<br>(Modified as per IDF criteria for South Asian, Chinese, and Japanese populations). | Male: $\geq 90$ cm<br>Female: $\geq 80$ cm |
| Triglyceride | $\geq 150$ mg/dl |
| HDL-C | Male: $<40$ mg/dL<br>Female: $<50$ mg/dL |
| Blood Pressure | $\geq 130/85$ mmHg or hypertensive on treatment. |
| Fasting Blood Sugar | $\geq 100$ mg/dl or diabetic on treatment |

#### Biochemical, Anthropometric, and Electrophysiological Parameters

**Table 2.** Biochemical, Anthropometric and Electrophysiological Parameters.

| **Parameters** | **Equipment** | **Test** | **Outcome Measure** |
| --- | --- | --- | --- |
| **Blood Pressure** | Aneroid<br>Sphygmomanometer | Auscultatory Blood Pressure<br>Monitoring | mmHg |
| **Fasting Blood Glucose** | Beck man coulter AU480<br>auto analyzer | Hexokinase method | mg/dL |
| **Postprandial Blood Glucose** | Beck man coulter AU480<br>auto analyzer | Hexokinase method | mg/dL |
| **HbA1C** | Tosoh G8HPLC analyzer | Ion exchange<br>High-performance liquid<br>chromatography | % |
| **Waist Circumference** | Inch Tape |  | cm |
| **Serum TAG** | Beck man coulter AU480<br>auto analyzer | Colorimetric assay by<br>coupled enzymatic reaction | mg/dL |
| **Serum HDL** | Beck man coulter AU480<br>auto analyzer | Colour-producing enzymatic<br>reaction | mg/dL |
| **Serum LDL** | Beck man coulter AU480<br>auto analyzer | Immunoprecipitation<br>followed by enzymatic<br>cholesterol measurement | mg/dL |
| **Heart rate Variability** | Powerlab, AD Instruments | Electrocardiography | - |

### Methodology

Participants were invited for short-term (5-minute) ECG recordings for HRV evaluation. Patients were telephonically reminded about the appointment and advised to wear loose clothes for the appointment, and to avoid heavy meals, caffeine, or smoking, and have a good night’s sleep one day prior. During the appointment, ECG leads were applied and the patients were rested for 20 minutes followed by short-term HRV testing.

### Statistical analysis

The data were maintained in Microsoft Excel 2019 and analyzed in IBM-SPSS Version 23. For HRV, only time and frequency domain parameters were analyzed. Spearman Correlation was done to study the association between MetScomponents and HRV parameters. With a confidence level of 95%, a p-value of < 0.05 was taken to be significant.

## Results

N=248 patients presented to the Internal Medicine Outpatient Department Lifestyle Disease Clinic were screened for MetS. Their anthropometric measures were recorded and routine blood investigations were sent. Of these, 174 individuals (n=87 males) were enrolled in the study. The descriptive statistics of metabolic syndrome components and HRV parameters among the study population are summarized in Table-3.

**Table-3.**
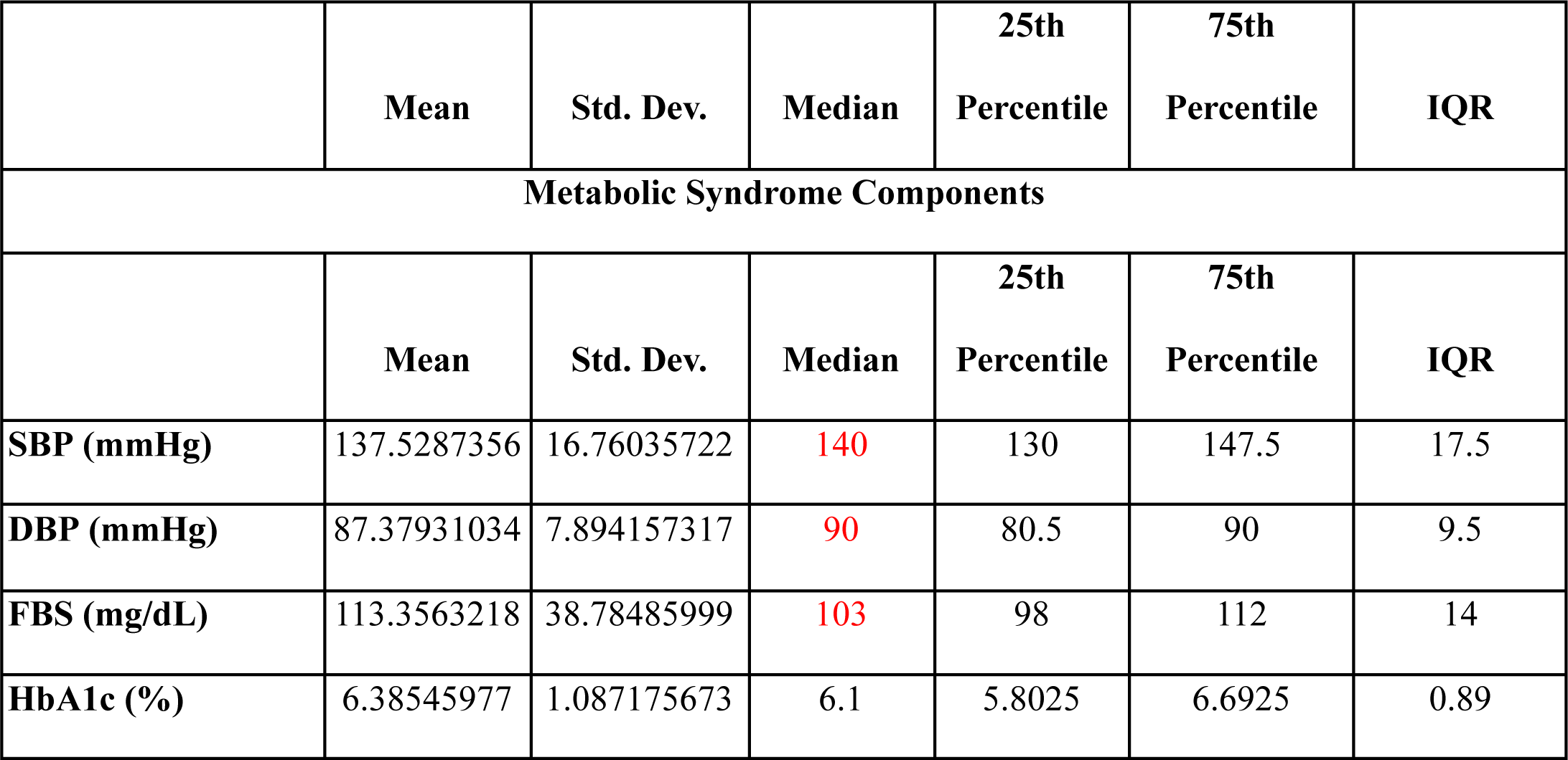
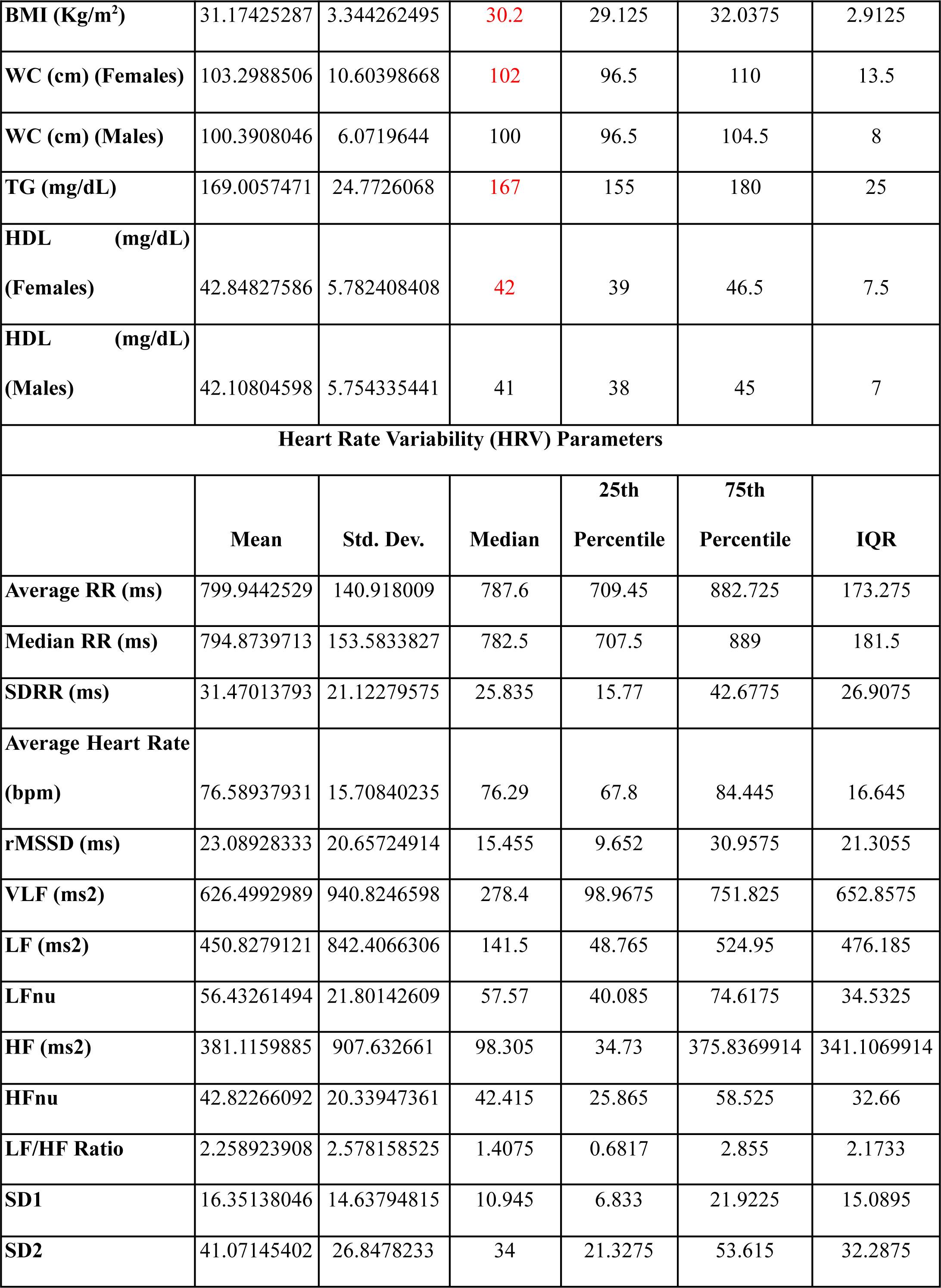
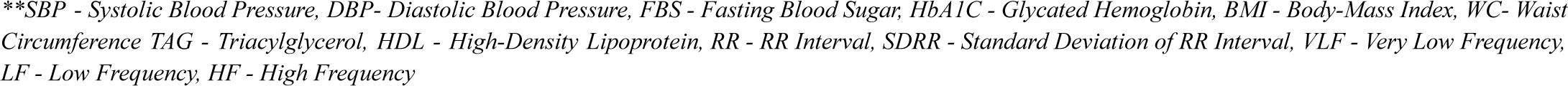
MetS components and HRV parameters of Metabolic Syndrome patients included in the study.

|  | Mean | Std. Dev. | Median | 25th Percentile | 75th Percentile | IQR |
| --- | --- | --- | --- | --- | --- | --- |
| **Metabolic Syndrome Components** |  |  |  |  |  |  |
|  | Mean | Std. Dev. | Median | 25th Percentile | 75th Percentile | IQR |
| **SBP (mmHg)** | 137.5287356 | 16.76035722 | 140 | 130 | 147.5 | 17.5 |
| **DBP (mmHg)** | 87.37931034 | 7.894157317 | 90 | 80.5 | 90 | 9.5 |
| **FBS (mg/dL)** | 113.3563218 | 38.78485999 | 103 | 98 | 112 | 14 |
| **HbA1c (%)** | 6.38545977 | 1.087175673 | 6.1 | 5.8025 | 6.6925 | 0.89 |
| **BMI (Kg/m<sup>2</sup>)** | 31.17425287 | 3.344262495 | 30.2 | 29.125 | 32.0375 | 2.9125 |
| **WC (cm) (Females)** | 103.2988506 | 10.60398668 | 102 | 96.5 | 110 | 13.5 |
| **WC (cm) (Males)** | 100.3908046 | 6.0719644 | 100 | 96.5 | 104.5 | 8 |
| **TG (mg/dL)** | 169.0057471 | 24.7726068 | 167 | 155 | 180 | 25 |
| **HDL (mg/dL) (Females)** | 42.84827586 | 5.782408408 | 42 | 39 | 46.5 | 7.5 |
| **HDL (mg/dL) (Males)** | 42.10804598 | 5.754335441 | 41 | 38 | 45 | 7 |
| **Heart Rate Variability (HRV) Parameters** |  |  |  |  |  |  |
|  | **Mean** | **Std. Dev.** | **Median** | **25th Percentile** | **75th Percentile** | **IQR** |
| **Average RR (ms)** | 799.9442529 | 140.918009 | 787.6 | 709.45 | 882.725 | 173.275 |
| **Median RR (ms)** | 794.8739713 | 153.5833827 | 782.5 | 707.5 | 889 | 181.5 |
| **SDRR (ms)** | 31.47013793 | 21.12279575 | 25.835 | 15.77 | 42.6775 | 26.9075 |
| **Average Heart Rate (bpm)** | 76.58937931 | 15.70840235 | 76.29 | 67.8 | 84.445 | 16.645 |
| **rMSSD (ms)** | 23.08928333 | 20.65724914 | 15.455 | 9.652 | 30.9575 | 21.3055 |
| **VLF (ms<sup>2</sup>)** | 626.4992989 | 940.8246598 | 278.4 | 98.9675 | 751.825 | 652.8575 |
| **LF (ms<sup>2</sup>)** | 450.8279121 | 842.4066306 | 141.5 | 48.765 | 524.95 | 476.185 |
| **LFnu** | 56.43261494 | 21.80142609 | 57.57 | 40.085 | 74.6175 | 34.5325 |
| **HF (ms<sup>2</sup>)** | 381.1159885 | 907.632661 | 98.305 | 34.73 | 375.8369914 | 341.1069914 |
| **HFnu** | 42.82266092 | 20.33947361 | 42.415 | 25.865 | 58.525 | 32.66 |
| **LF/HF Ratio** | 2.258923908 | 2.578158525 | 1.4075 | 0.6817 | 2.855 | 2.1733 |
| **SD1** | 16.35138046 | 14.63794815 | 10.945 | 6.833 | 21.9225 | 15.0895 |
| **SD2** | 41.07145402 | 26.8478233 | 34 | 21.3275 | 53.615 | 32.2875 |

*\*\*SBP - Systolic Blood Pressure, DBP- Diastolic Blood Pressure, FBS - Fasting Blood Sugar, HbA1C - Glycated Hemoglobin, BMI - Body-Mass Index, WC- Waist Circumference TAG - Triacylglycerol, HDL - High-Density Lipoprotein, RR - RR Interval, SDRR - Standard Deviation of RR Interval, VLF - Very Low Frequency, LF - Low Frequency, HF - High Frequency*

The anthropometric and biochemical parameters were then tested for any significant correlation with HRV parameters as shown in Table-4.

**Table-4.** Spearman test between MetS components and HRV parameters.

|  | **SBP** | **DBP** | **WC** | **FBS** | **TG** | **HDL** |
| --- | --- | --- | --- | --- | --- | --- |
| **AVG HR** | 0.030<br>(0.74) | 0.030<br>(0.67) | -0.01<br>(0.93) | 0.12<br>(0.13) | 0.12<br>(0.129) | 0.090<br>(0.21) |
| **AVG RR** | -0.02<br>(0.77) | -0.03<br>(0.65) | -0.17<br>**(0.02)** | 0.03<br>0.65 | -0.09<br>(0.22) | -0.1<br>(0.18) |
| **SDRR** | 0.02<br>(0.79) | 0<br>(0.96) | -0.08<br>(0.28) | 0.09<br>(0.24) | -0.07<br>(0.34) | 0.08<br>0.314 |
| **rMSSD** | -0.05<br>(0.520) | -0.03<br>(0.702) | -0.23<br>**(0.002)** | 0.08<br>(0.319) | -0.06<br>(0.400) | 0.05<br>(0.49) |
| **VLF** | 0.09<br>(0.221) | 0.07<br>(0.385) | -0.04<br>(0.597) | 0.02<br>(0.806) | -0.11<br>(0.159) | 0.07<br>(0.376) |
| **LF** | 0<br>(0.955) | -0.03<br>(0.721) | 0.03<br>(0.704) | 0.02<br>(0.841) | -0.02<br>(0.804) | 0.1<br>(0.184) |
| **LFnu** | 0.04<br>(0.559) | -0.05<br>(0.492) | 0.49<br>**<0.001** | 0.02<br>(0.843) | 0.15<br>(0.53) | 0.1<br>(0.188) |
| **HF** | -0.04<br>(0.596) | -0.05<br>(0.475) | -0.22<br>**(0.003)** | 0.04<br>(0.562) | -0.08<br>(0.284) | 0.04<br>(0.557) |
| **HFnu** | -0.05<br>(0.555) | 0.05<br>(0.539) | -0.49<br>**<0.001** | -0.01<br>(0.896) | -0.15<br>(0.051) | -0.11<br>(0.162) |
| **LF/HF** | 0.04<br>(0.622) | -0.05<br>(0.494) | 0.49<br>**<0.001** | 0.02<br>(0.82) | 0.15<br>**(0.048)** | 0.11<br>(0.165) |

Waist circumference (WC) was reported to have a significant moderate to strong positive correlation with sympathetic predominant HRV parameters - LFnu and LF/HF ratio. (Table-4) Parasympathetically modulated HRV parameters like Average RR, rMSSD, and HF had a negative correlation with WC. (Table-4). No other biochemical or anthropometric measure shows a consistent correlation to autonomic parameters. (Table-4) The significant correlations have been described in greater detail in Table-5.

**Table-5.**
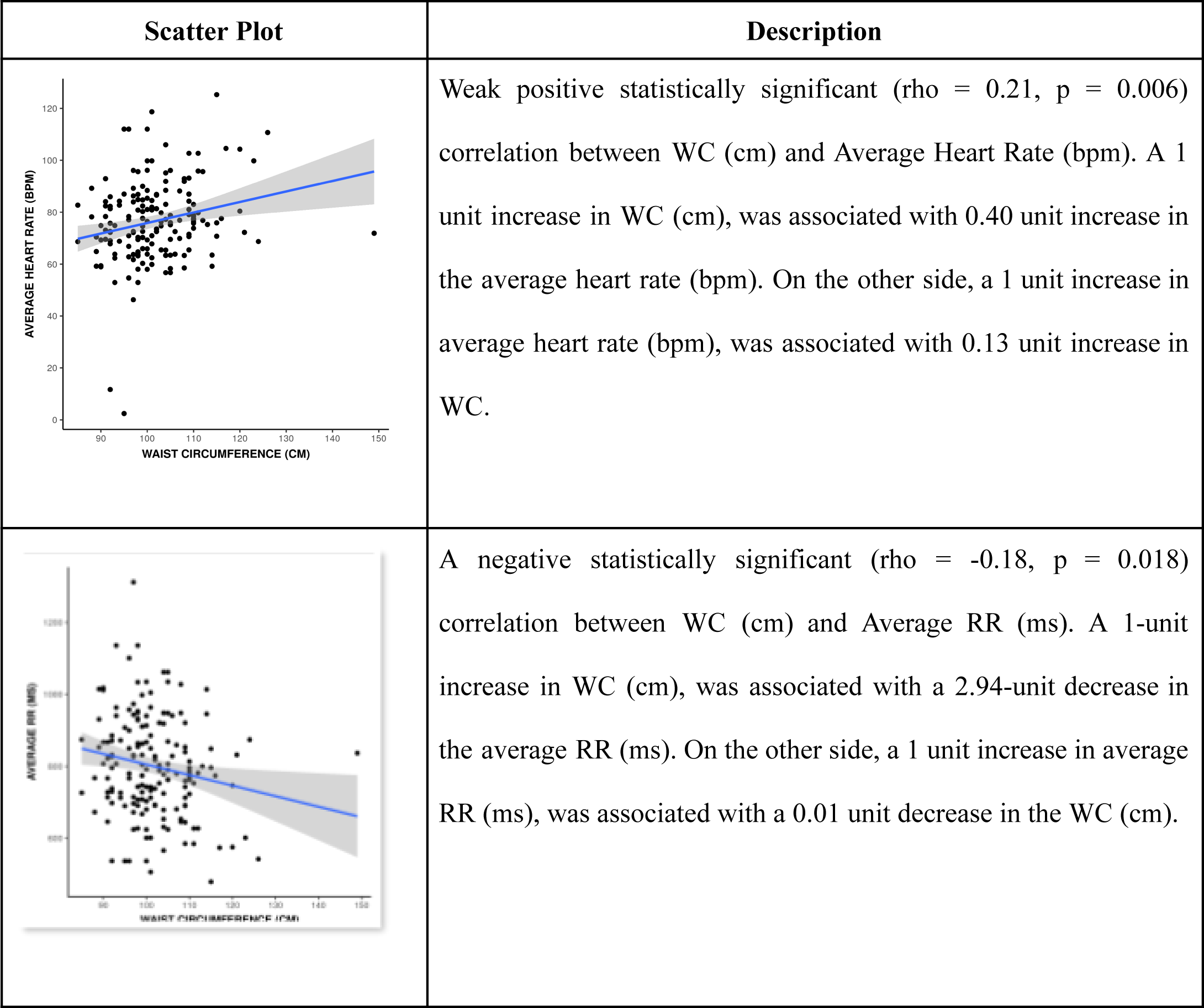
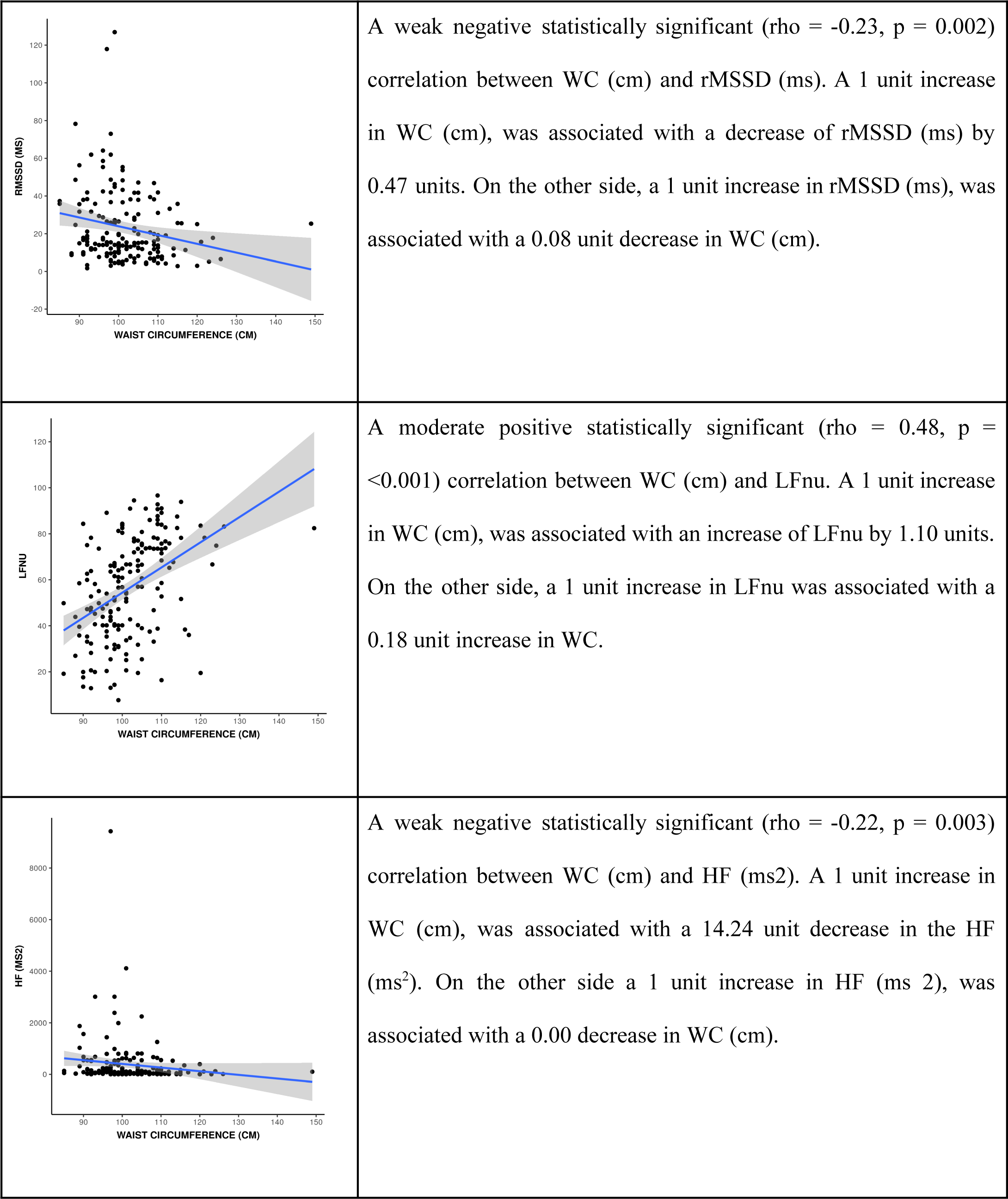
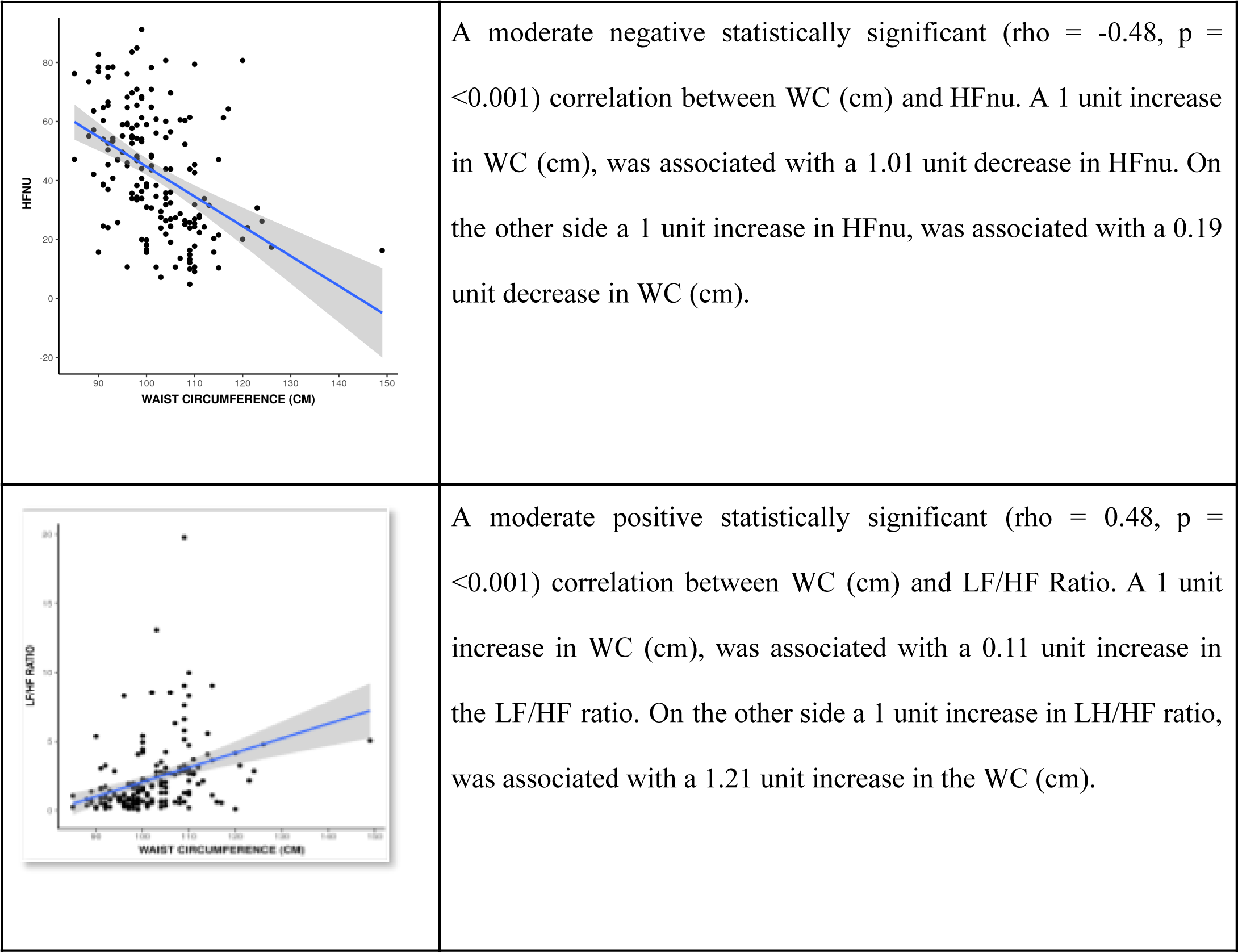
Scatter plots depicting significant correlations between Autonomic parameters, and waist circumference.

## Discussion

HRV is a validated cAFT known to record the general and especially the cardiac autonomic profile accurately. (27) While, it is considered ideal to have a 24-hour ECG recording for accurate evaluation of HRV, short-term (5-minute) ECG recordings, have long been a convenient tool to reliably study cardiac autonomic function. (27) ECG can be analyzed to generate time-domain, frequency-domain, and non-linear measurements. Time-domain HRV parameters are Avg HR, Avg RR, SDRR, SDANN, SDNN Index, RMSSD, NN50, and pNN50 of which Avg HR, Avg RR, SDRR, and RMSSD were analyzed in the current study. Of the frequency-domain parameters, VLF, LF, and HF frequency parameters were analyzed. Among the analyzed parameters, RMSSD, and HF nu are well-known indicators of parasympathetic activity, and LF nu and LF/HF ratio, of sympathetic activity. (27)

In Shaffer F. et. al.’s words, the autonomic nervous system must not be understood as a “zero-sum” combination of sympathetic and parasympathetic nervous activity. (27) Hence, it is ideal to discuss the sympathetic and parasympathetic nervous activity separately. The primary autonomic derangement reported in MetS is a general sympathetic overdrive. (15, 30-32) Evidence from a variety of experiments and studies has been reporting the same for over a decade. (30, 31) This sympathetic overdrive at the level of the heart plays a pathogenic role in cardiovascular comorbidities associated with MetS. (15) The profile of parasympathetic activity still has scope to be better understood, although studies have reported a loss of recovery of heart rate after a stressful situation, (a cardiac parasympathetic function) to be impaired in the setting of MetS. (33, 34) In toto, cardiac sympathetic activity increases while cardiac parasympathetic activity decreases in the setting of MetS and poses a cardiovascular risk.

Several attempts have been made to quantify these cardiac autonomic imbalances in MetS through cAFTs, and hence predict cardiovascular risk based on these results. HRV parameters reported to be significantly deranged in MetS are rMSSD, LF power, HF power, and LF/HF ratio. (35–39) The cardiac sympathetic overdrive in the setting of MetS manifests as an elevated LF power/LF nu and a high LF/HF ratio. (35–39). The HF power/HF nu has been reported to decrease in MetS. (36–39) These findings pose a cardiovascular risk in several pathogenic backgrounds including MetS over the long term.

These findings from the literature enable us to streamline our approach toward identifying an anthropometric or biochemical proxy for the cardiac autonomic profile in MetS. A given biochemical or anthropometric parameter having moderate to strong statistically significant correlation with the HF, LF, and LF/HF ratio might be a suitable proxy. In the current study, only WC showed a significant (p<0.05) moderate (r=0.49) correlation with LF nu, HF nu, and LF/HF ratio. The nature of the correlation of WC with LF nu and LF/HF ratio was positive while that with HF nu was negative. This suggests that WC might be used as a proxy for the cardiac autonomic profile in the setting of MetS.

Similar to our study, a recent study from Southern India analyzed the correlation between anthropometric/biochemical parameters and cardiac autonomic function among pre-MetS patients and reported a significant but weak correlation between abdominal obesity or waist-hip ratio, and LF/HF ratio. (37) A meta-analysis summarising data published until 2014 reported a strong correlation between WC and cardiac autonomic function parameters. (40) It further emphasizes this correlation to be stronger than association with any other anthropometric or biochemical parameter. (40) Original work led by the same author in the following year also reported WC to be most commonly associated with HRV (41) A meta-analysis studying similar objectives among the pediatric population reported a significant correlation between WC and LF/HF ratio and HF nu after an analysis of 3,000+ patients each. (42) These results can form the basis for validation of WC as an indicator of cardiac autonomic profile in MetS in rural and resource-limited healthcare setups. While HRV analysis requires an ECG machine, algorithms/software in place to analyze HRV, and trained medical staff to understand and interpret the results, WC can be measured using an inch tape available in every household. Moreover, it does not require much technical expertise in analyzing or interpreting it. Although a parameter like WC is more likely to vary across populations and cultures, thus it requires significant effort to establish valid cut-offs especially if it is to be used as an indicator of cardiac autonomic profile. Hence, WC might be a measure with significant practical application in primary and community healthcare.

It is worth noting that, WC has been the representative anthropometric measure of abdominal obesity and visceral adiposity for decades. The expanding understanding of the etiopathogenesis of MetS has led to the conclusion that of the four diagnostic components of MetS, the causative role of visceral adiposity is the most well-established. (43) Visceral adiposity has been shown to trigger most pathways of MetS. (43–45) Moreover, adipose hypertrophy even in the absence of confounding insulin resistance or Type-2 Diabetes Mellitus, atherosclerotic cardiovascular disease, smoking, macro-inflammation, or lipid therapy has been shown to be associated with MetS. (46) Visceral adiposity increases the supply of free fatty acids, especially in the splanchnic circulation, and hence to the liver in turn contributing to insulin resistance. (43, 47) Insulin resistance underlies the remaining three diagnostic components of MetS - elevated blood glucose, dyslipidemia, and hypertension at least partly. (43, 48) This hypothesis of visceral adiposity being the induction point of MetS provides a logical explanation behind WC being a suitable indicator of cardiac autonomic function. Since visceral adiposity is present at the scene of crime since (or maybe even before) the precipitation of MetS, its anthropometric indicator (WC), and complication (namely cardiac autonomic imbalance) are likely to be correlated in the disease setting.

The meta-analysis from 2014 reported an inconsistent relation between cardiac autonomic function and blood pressure as well. (40) The associations were significant among children but not among adults. (40) Just like blood pressure, inconsistent associations were also reported between cardiac autonomic function and fasting plasma glucose, and triglycerides. (40) Contrary to the current study, there are reports of hyperglycemia also being a reliable predictor of cardiac autonomic function. (49, 50) The meta-analysis of the pediatric population reported different cardiac autonomic parameters to have significant correlations with SBP, DBP, WC, TAGs, and HDL. (42) There is a major inconsistency in the literature related to correlations between cardiac autonomic function and MetS parameters. A number of hypotheses can be postulated to understand the difference between the findings of the current study and those published in the literature. The vast pathophysiological background of metabolic syndrome creates scope for anthropometric and biochemical parameters of the disease to be affected by an even larger number of variations in diet, lifestyle, and day-to-day stress to name a few. These factors definitely differ between individuals and even more significantly between populations. Similarly, inherited and acquired genetic mutations may also play a role in the same disease expressing a different biochemical or anthropometric phenotype. Such variation may amount to the inconsistency discussed above.

Hence, it is safe to say that the current study identifies WC as a potential indicator of cardiac autonomic profile in MetS most reliably for residents of western Uttar Pradesh and Uttarakhand. It also emphasizes the need for center-focused studies to identify population-specific proxies for diagnostics like HRV to facilitate public and community healthcare.

Validation of population-specific WC cut-offs and WC-based risk stratification by employing alternate tools might be the steps ahead to understand the relation between WC and cardiac autonomic function or cardiovascular risk. A longitudinal study following up the biochemical and anthropometric measures of patients throughout the course of the disease might give us more information. Also, a better lifestyle, socio-cultural, and genetic profiling of MetS patients might reveal different phenotypic variations of the disease and hence yield information on the correlative incongruency identified in the existing literature.

## Conclusions

The study identifies WC as a suitable indicator for cardiac autonomic profile in patients with MetS and by extension of cardiovascular posed by the disease. WC is of particular use for a country with limited healthcare resources and hence must be further probed into for its mass-scale practical applications.

## Conflict of Interest

The authors do not have any conflict of interest to declare.

## External Funding

The study was conducted at All India Institute of Medical Sciences (AIIMS), Rishikesh, Uttarakhand, India. No external funding was applied for or received for the conduction of this study.

## Data Availability

All data produced in the present study are available upon reasonable request to the authors.

## Notes

### Competing Interest Statement

The authors have declared no competing interest.

### Funding Statement

This study did not receive any funding.

### Author Declarations

Ethics Committee of All India Institute of Medical Sciences (AIIMS), Rishikesh, Uttarakhand, India (DHR Reg: EC/NEW/Inst/2020/1046 CDSCO Reg: ECR/736/Inst/UK/2015/RR-21) gave ethical approval for this work.

